# Maternal Metabolomic Signatures of Spontaneous Preterm Birth in Bangladesh: Evidence from the AMANHI Bangladesh Cohort

**DOI:** 10.1101/2025.08.07.25333197

**Authors:** Bipasha Akhter, Radiah Azmyne Khan, Md Shafiqul Islam, Rasheda Khanam, Nabidul Haque Chowdhury, Salahuddin Ahmed, Tarik Hasan, Kim Lagerborg, Tao Long, Cameron Lamoureux, Muhamad Maqsud Hossain, Nilanjan Chatterjee, Abdullah H. Baqui, Ni Zhao

## Abstract

Spontaneous preterm birth (sPTB), defined as natural labor onset before 37 weeks of gestation, is a major contributor to neonatal morbidity and mortality, especially in low- and middle-income countries. Despite its global impact, the biological mechanisms underlying sPTB remain poorly understood. This study employed untargeted metabolomics and a nested case-control study design to investigate maternal serum metabolites that are differentially expressed in sPTB and the associated biological pathways. Cases (sPTB, n=267) and controls (healthy term births, n=864) were drawn from a prospective pregnancy cohort of 3,000 women enrolled in the Alliance for Maternal and Newborn Health Improvement (AMANHI) study in Bangladesh. Study participants were selected in early pregnancy from two rural subdistricts of the Sylhet district in Bangladesh, with data collected between 2014 and 2018. Maternal blood samples were obtained at two time points during pregnancy: once in early pregnancy (before 20 weeks) and in late pregnancy (at or after 20 weeks). Serum metabolites were profiled using an untargeted metabolomics approach based on high-throughput untargeted liquid chromatography-mass spectrometry (LC-MS), yielding 55,541 metabolites, for which 650 were annotated. Statistical analyses revealed significant associations (FDR < 0.1) between sPTB and 268 metabolites in early pregnancy and 617 in late pregnancy. In cases of sPTB, triglyceride levels were markedly reduced in early pregnancy. In contrast, phosphocholines, sphingomyelins, and very long-chain dicarboxylic acids exhibited differential expression in late pregnancy, suggesting disruptions in lipid metabolism. The rate of change in hydroxyisovaleric acid levels across paired samples was significantly higher in sPTB cases, indicating its potential as a dynamic biomarker. Metabolite set enrichment analyses highlighted key differences in triglyceride and dicarboxylic acid pathways, implicating roles in energy metabolism, inflammation, and placental function. Dihydrothymine was elevated across both time-points, indicating its role in pyrimidine metabolism. These findings provide valuable insights into the biological basis of sPTB and highlight the potential of metabolomic biomarkers for risk assessment and targeted intervention.

## Introduction

Spontaneous preterm birth (sPTB), which accounts for approximately 70% of all PTB cases, is defined as childbirth before 37 weeks of gestation and is initiated by either the spontaneous onset of labor or preterm premature rupture of membranes (pPROM) (1,2). Preterm birth is a leading cause of neonatal morbidity and mortality worldwide, responsible for over 1 million <5 deaths each year. In addition, PTB substantially contributes to long-term health complications among survivors, manifesting in significant respiratory, neurological, and developmental disorders (3,4). In 2020, about 13.4 million newborns, accounting for 9.9% of total births, were born preterm (5). Despite advancements in antenatal care, the incidence of sPTB remains high, particularly in low- and middle-income countries (6). Notably, between 70% and 80% of global preterm births occur in Southern Asia and sub-Saharan Africa. Within South Asian, Bangladesh had the highest rate of preterm births at 16.2% in 2020, whereas our group has documented a rate of 13.2% (7,8).

PTB is complex, with unclear and multifactorial etiology. Thus, effective screening of women with high risk of PTB can be challenging (9). Several mechanisms have been linked to PTB, including neuroendocrine, vascular, immune-inflammatory, and behavioral factors. Research has examined key markers relating to uterine distension, decidual inflammation or infection, and activation of the hypothalamic-pituitary-adrenal axis in the context of sPTB. However, neither individual markers nor combinations thereof have proven reliably accurate for the prediction of sPTB. While clinical indicators like previous PTB history, second-trimester cervical length, and cervico-vaginal fetal fibronectin (fFN) demonstrate some potential, they generally are not useful for asymptomatic women and only predict sPTB at later pregnancy stages (1,10). Different pathways may contribute to the development of sPTB at different stages of pregnancy; however, reliable biochemical tools for accurately predicting it at each stage are still lacking (11,12).

Given the complex etiology of sPTB, metabolomics emerges as a particularly promising field. By offering comprehensive biochemical profiles, this approach is uniquely positioned to address the limitations of previous studies by detecting subtle metabolic alterations that precede sPTB (13). Unlike other omics technologies, metabolomics provides a direct snapshot of cellular activity because it measures the end products of cellular processes (14). The untargeted metabolomics approach, which simultaneously assesses all metabolites in a sample, is a robust and unbiased approach for studying sPTB. By identifying and relatively quantifying thousands of metabolites simultaneously, untargeted metabolomics could enhance biomarker discovery and advance understanding of physiology and disease (15). This comprehensive method holds considerable potential for uncovering the metabolic underpinnings of sPTB, offering insights that could lead to more effective predictive tools and therapeutic strategies (9).

Despite the high burden of spontaneous preterm birth (sPTB) in low- and middle-income countries (LMICs), omics-based research—including metabolomics - remains limited in these settings, making this study the first untargeted metabolomics approach on a large cohort of pregnant women in an LMIC like Bangladesh (16). Moreover, while most published metabolomics studies focus on static biomarkers, our use of paired early and late pregnancy samples enables longitudinal metabolomic profiling within the same individuals, allowing us to capture dynamic changes over time and identify gestational-age specific biomarkers and those more closely linked to the progression of sPTB (17). Finally, as the future direction of metabolomics research moves beyond identifying individual differential metabolites, there is growing emphasis on understanding biological pathways and causal mechanisms (18). By integrating metabolite profiling with metabolite set enrichment analysis, our study aims to also uncover and understand the processes underlying the metabolomic signatures and their potential contribution to sPTB.

## Methods

### Study Design

#### Setting

We conducted a nested case-control study utilizing data from the Alliance for Maternal and Newborn Health Improvement cohort at Bangladesh site (AMANHI-Bangladesh), a population-based study of pregnant women. AMANHI-Bangladesh is part of the MOMI Consortium, an international collaborative initiative that integrates multi-omics data from multiple maternal and infant cohorts, including AMANHI, to advance the understanding of biological mechanisms underlying maternal and neonatal health outcomes. As part of the AMANHI-Bangladesh, a comprehensive biorepository was established, comprising data and biospecimens from 3,000 pregnant women recruited between 21 August 2014 and 30 Aug 2017 across two rural sub-districts in the Sylhet district. Pregnancies were systematically identified through two-monthly home visits, confirmed via strip-based pregnancy tests, and gestational age (GA) was determined using ultrasonography between 8–19 weeks of gestation. Community health workers conducted three structured antenatal visits (8–19, 24–28, and 32–36 weeks of gestation) and two postnatal visits (< 7 and 42–60 days postpartum), collecting extensive phenotypic, socio-demographic, and epidemiological data (8,19). Written informed consent was obtained from all mothers in the cohort. For children, written informed consent was obtained from a parent or legal guardian.

#### Sample Collection

Maternal blood samples were collected from pregnant women in the cohort by trained phlebotomists at 8–19 weeks GA, with a second sample collected at 24–28 GA or at 32–36 weeks. Samples were processed following standard protocol, including centrifugation to separate serum and plasma, aliquoting and storage −80°C (19).

#### Participant Selection

From the initial cohort of 3,000 pregnant women, we excluded participants if metabolomics data were unavailable. Further exclusion criteria include non-singleton pregnancies, missing gestational age data, stillbirths, or induced labor. After applying the exclusions, a case-control study was conducted, where cases comprised women who experienced sPTB. Controls were selected from women who had healthy term births (39–41 weeks of gestation) and were further restricted to exclude conditions such as low birth weight, chronic hypertension, diabetes, congenital anomalies, or maternal infections.

### Metabolomics Analysis

#### Sample Processing

Human plasma samples are thawed over 5 minutes using Sapient’s rapid plate thaw system and placed on an orbital shaker at 550 rpm at 4°C for 10 minutes. A 16 µL aliquot of plasma is then transferred to a shallow well 96-well microtiter plate containing 200 µL of extraction solution [methanol:acetonitrile (35:65) with 0.5% formic acid, containing the following internal standards: ^13^C ^15^N -glutamate (Sigma-Aldrich), CUDA (Cayman Chemicals), arachidonic acid-d11 (Avanti Lipids), PC(18:1/16:0-d31) (Avanti Lipids), TG(14:0/16:1/14:0-d5) (Avanti Lipids)]. Samples are then shaken at 700 rpm at 4°C for 10 minutes followed by centrifugation at 6000g at 4°C for 10 minutes after which 2 mL (Pos mode) or 15 mL (Neg mode) of supernatant was transferred to a 384-well glass-lined polypropylene plate containing either 148 mL 20:80 methanol:water + 1 mM ammonium formate and 0.05% formic acid or 135 mL 40:60 methanol:water + 0.1% acetic acid (for Pos and Neg mode analysis, respectively) each containing additional internal standards: phenylalanine-d6 (Cayman Chem) and MAPCHO-12-d38 (Avanti Lipids). In total 7 internal standards across positive and negative electrospray ionization modes are monitored for system performance including monitoring sample stability, additive effects, matrix effects and column performance.

#### rLC-MS Data Acquisition

rLC-MS based metabolomics measures are performed using Sapient’s next generation, high-throughput, rapid liquid chromatography-mass spectrometry (rLC-MS) technology. Sapient’s rLC-MS consists of a proprietary chromatographic system coupled to a high-resolution Bruker timsTOF Pro 2 mass spectrometer (Bruker). Samples undergo a 52 second chromatographic separation using a custom packed mixed-mode column for data acquisition in both positive and negative electrospray ionization modes using mobile phases consisting of water, methanol, isopropanol, acetic acid and ammonium acetate. A separate aliquot of pooled commercial plasma is also extracted and injected as an external QC sample about every 30 injections. Mass spectrometer parameters are set according to the following: dry gas temp of 350 °C, dry gas flow rate of 8 L/min, nebulizer gas of 50 psi, probe gas temp of 150(+)/250(-) °C, probe gas flow rate of 3 L/min, capillary voltage of 3750 V and End Plate voltage of (+) 2000 / (-) 1500 V, mass range set to 50 – 1700 m/z, with data acquisition at 9 spectra/second.

#### Spectral Data Handling, Alignment & Standards Matching

Raw rLC-MS data files are converted to mzXML using MSConvert (Proteowizard software package). Chromatographic drift is corrected according to landmark based spectral alignment as we have previously described (20). Metabolite features are extracted from drift-corrected mzXML files using custom neural network-based software (21). Following data extraction, metabolite features are normalized using a batch median normalization method with correction for median levels. rLC-MS analysis of human plasma samples typically enables measurement of 11,000-15,000 small molecule metabolites per biospecimen.

#### Quality Assessment of Spectral data

QC/QA of rLC-MS data is performed using the panel of isotopically labeled internal standards as well as bracketed pooled plasma samples to monitor fluctuations in extraction efficiency, instrument sensitivity, matrix artifact and mass accuracy. For system suitability, mass calibration is performed every 24 hours and used to assess mass accuracy, mass resolution, detector sensitivity, and instrument cleanliness. For each 384-well plate run, QC assessment consists of (i) isotopically labeled internal standards added to each sample at the first sample preparation step and the supernatant dilution step to monitor sample to sample matrix effects, (ii) bulk pre-aliquoted commercial pooled plasma placed in wells A1, D12 and H12 of each 96-well sample plate and prepared identically to human samples (internal bracket QC sample), (iii) bulk pre-aliquoted commercial pooled plasma prepared external to samples by hand (external bracket QC sample), and (iv) a preparation blank prepared during each 384-well plate to assess background. Overall coefficient of variation (%CV) is be monitored for internal standards in all samples. For samples with %CV >25% and total ion current measures >3 standard deviations from mean, samples undergo re-injection.

#### Compound Identification

Initial compound identification is performed from MS1 experimental data whereby accurate mass (+/- 10ppm) and retention time (+/- 0.015 minutes) is matched against Sapient’s Global Identification database. This manually curated database contains only signals previously observed and annotated from biological samples and confirmed by accurate mass, tandem-MS fragmentation pattern, ion mobility (CCS) value and retention time (when a commercial standard was available). Furthermore, statistically prioritized signals are confirmed through targeted tandem-MS data collection and/or injection of commercially available standards alongside client biosamples. For unknown compounds, each prioritized feature is manually inspected to confirm chromatographic peak quality and to ensure each feature is the monoisotopic MS1 peak for its peak cluster. Tandem-MS spectra are then collected for each feature and searched against Sapient’s chemical data repository, which contains experimental and in-silico MS2 for over 150k compounds. Unmatched MS2 spectra are then manually inspected and when possible annotated based on the presence of diagnostic MS2 fragments or diagnostic neutral losses.

#### Batch normalization

All biosamples were run in 384-plates on Sapient’s rLC-MS platform. To account for variations among plates and to avoid overfitting of the data, a simple batch normalization algorithm has been applied to yield the extracted metabolite feature intensities:

#### Metabolomics data imputation

Missing metabolite intensities were imputed based on assumptions of below-the-limit-of-detection missingness. Particularly, for each metabolite feature, its missing values in samples were filled in with the values randomly drawn from a uniform distribution range from min/10 to min, where min is the minimum intensity of the feature among all samples. Metabolite levels were then log2 transformed followed by standardization per metabolite by subtracting the mean and scaling to unit variance with the standard score of metabolite feature calculated as z = (x-u) / s where u is the mean and s is the standard deviation of the metabolite abundances across samples.

### Statistical Data Analysis

#### Participant Baseline Characteristics

Maternal and household characteristics were summarized as frequencies and percentages for sPTB and control groups. Associations between these variables and sPTB status were assessed via Chi-square tests, and at p value < 0.05 was considered statistically significant.

#### Principal Component Analysis and Metabolomic Variation

Principal Component Analysis (PCA) was conducted on the metabolomics data for dimension reduction and visualization, separately for early (< 20 weeks GA) and late (≥ 20 weeks GA) pregnancy samples. Global association between metabolomics profiles and sPTB status was assessed via Permutational Multivariate Analysis of Variance (PERMANOVA) with 999 permutations, stratified by gestational age groups (22).

#### Metabolite-Wise Association Analyses

Logistic regression models were applied to each of the 55,541 metabolites (both annotated and non-annotated) to identify metabolites associated with sPTB.

The analysis was separated into three logistic regression models to examine how maternal metabolite levels are related to sPTB. The first model looked at metabolite levels in early pregnancy samples (< 20 weeks GA), and the second model looked at levels in late pregnancy samples (≥ 20 weeks GA). The third model focused on changes in metabolite levels over time, using paired samples from the same mother. For each metabolite, we calculated the weekly rate of change by subtracting the intensity at the early time point from the intensity at the late time point and dividing by the number of weeks between the two samples. This model was adjusted for the baseline (early pregnancy) metabolite level to control for individual differences in starting levels. Maternal age was used as a covariate for adjusting all three regression models as it is an established potential confounder of preterm birth (23). Statistical significance was determined at the Benjamini-Hochberg false discovery rate (FDR) threshold of < 0.1(24). By using these three models, we aimed to identify both static and dynamic metabolic markers associated with sPTB.

#### Pathway Enrichment Analysis

Pathway enrichment analysis was performed using Fast Gene Set Enrichment Analysis (FGSEA) on annotated metabolites from (< 20 weeks GA) and late (≥ 20 weeks GA) pregnancy samples separately (25). Metabolites were categorized into distinct compound classes, and enrichment analysis was conducted to assess their functional relevance. For FGSEA, signed p-values derived from individual logistic regression models were used to rank all annotated metabolites, and the FGSEA algorithm was then applied to compute enrichment scores for each compound class (26).

#### Software and Reproducibility

All statistical analyses were conducted on R version 4.3.3. Analysis code and data files, R packages necessary to reproduce the analyses are available upon request.

#### Ethics Approval and Consent to Participate

The study protocol received ethical approval from the following committees: the Institutional Review Board of icddr,b (PR12073, approved on 23 March 2014), the Johns Hopkins Bloomberg School of Public Health (IRB 00004508, approved on 8 August 2012), and the World Health Organization (RPC 532, approved on 22 July 2014). Informed consent was obtained from all participants and/or their legal guardians.

## Results

### Participant Selection and Baseline Characteristics

From the initial cohort of 3,000 pregnant women, a total of 1,131 participants were included in this metabolomic case-control study, comprising 267 sPTB cases (live births <37 weeks) and 864 term controls (births at 39–41 weeks). Fig 1 provides a detailed overview of participant exclusions based on predefined criteria.

**Fig 1:**
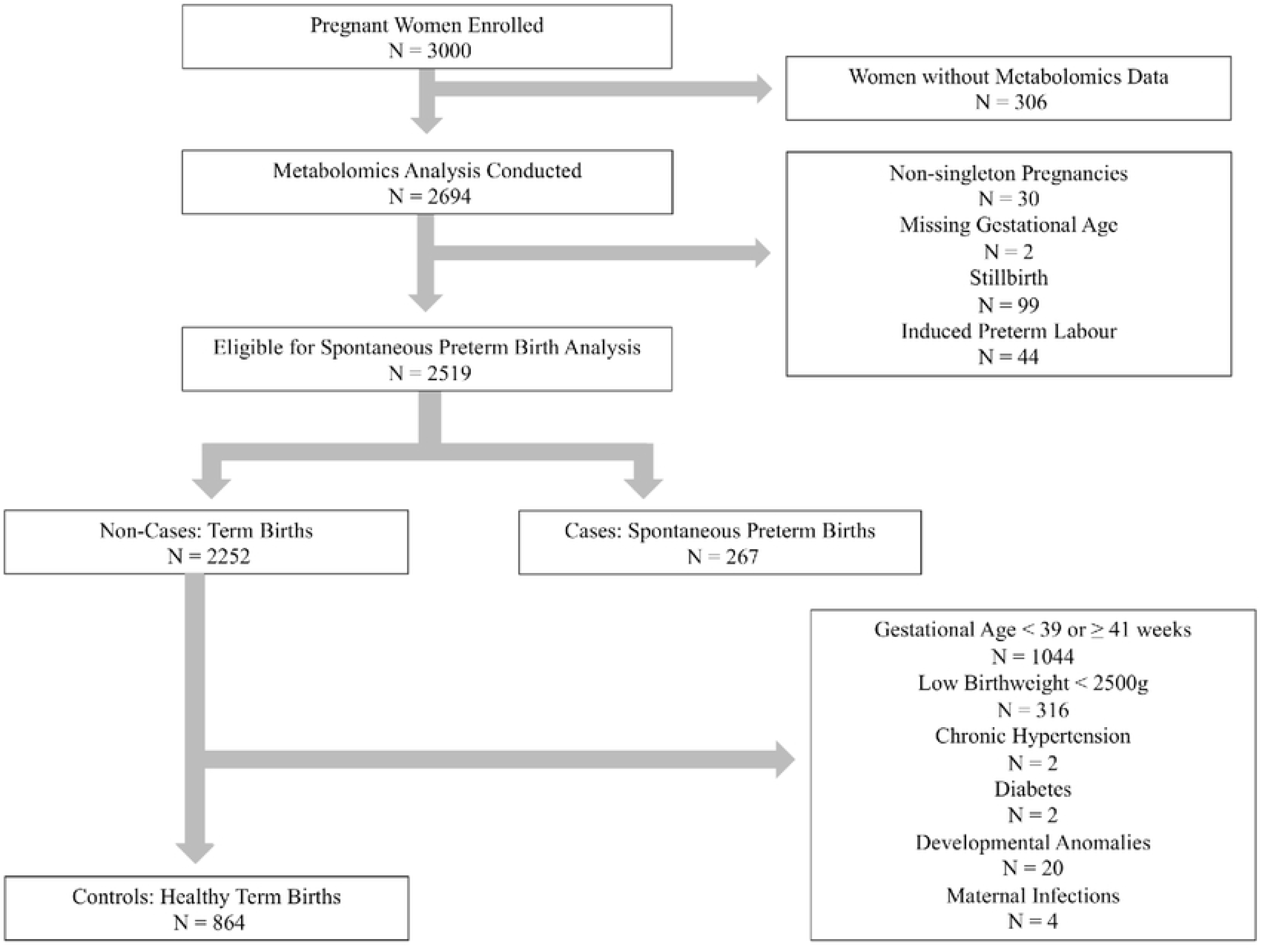
Flowchart of Study Participants Selection.

Women in our cases group with sPTB had significantly shorter maternal height and lower early pregnancy BMI compared to controls. Additionally, a higher proportion of women in the sPTB group were nulliparous (p < 0.001) and had husbands with ≤ 5 years of education (p < 0.001). A significant association was also observed between women’s occupation and sPTB status (p = 0.02); however, the number of women who were employed and worked outside the home was relatively small. No significant differences were observed between sPTB and control in maternal age groups or tobacco sniffing/chewing habits or passive smoke exposure. A detailed summary of these demographics characteristics of the study participants can be found in Table 1.

**Table 1:** Maternal and Household Characteristics and their Association with Spontaneous Preterm Birth (sPTB)

| Maternal Characteristics | sPTB<br>N= 267 (23.6%) | Control<br>N=864 (76.4%) | P. value |
| --- | --- | --- | --- |
| Maternal Age group |  |  |  |
| <30 yrs | 238 (23.9%) | 758(76.1%) | 0.61 |
| >=30 yrs | 29 (21.5%) | 106(78.5%) |  |
| Mother's Height |  |  |  |
| <145 cm | 65(35.9%) | 116(64.1%) | < 0.001 |
| >=145 cm | 202(21.3%) | 748(78.7%) |  |
| Mother's Early Pregnancy BMI |  |  |  |
| Underweight | 114(30.7%) | 257(69.3%) | < 0.001 |
| Normal Weight | 142(20.7%) | 545(79.3%) |  |
| Overweight/Obese | 11(15.1%) | 62(84.9%) |  |
| Parity |  |  |  |
| Nulliparous | 97(29.6%) | 231(70.4%) | < 0.001 |
| 1-3 | 139(20.7%) | 531(79.3%) |  |
| 4 or more | 31(23.3%) | 102(76.7%) |  |
| Mother's Occupation |  |  |  |
| Housewife | 260(23.3%) | 858(76.7%) | 0.02 |
| Employed | 7(53.8%) | 6(46.2%) |  |
| Husband's Education |  |  |  |
| ≤ 5 years | 204(27.6%) | 536(72.4%) | < 0.001 |
| > 5 years | 63(16.1%) | 328(83.9%) |  |
| Tobacco sniffing/chewing |  |  |  |
| Never | 215(23%) | 719(77%) | 0.4 |
| Quit during pregnancy | 1(33.3%) | 2(66.7%) |  |
| Currently sniffing/chewing | 51(26.3%) | 143(73.7%) |  |
| Passive Smoking Exposure |  |  |  |
| Not Exposed | 50 (21.5%) | 183 (78.5%) | 0.4 |
| Exposed | 217 (24.2%) | 681 (75.8%) |  |

### Principal Component Analysis and Metabolomic Variation

Fig 2 displays the first two principal components (PCs) of the metabolite profiles, with points colored by sPTB status and pregnancy stage (early or late pregnancy). There is a clear difference between metabolite profiles from early and late pregnancy samples. However, when stratified by early or late pregnancy, no significant differences were detected in metabolite profiles based on sPTB status.

**Fig 2:**
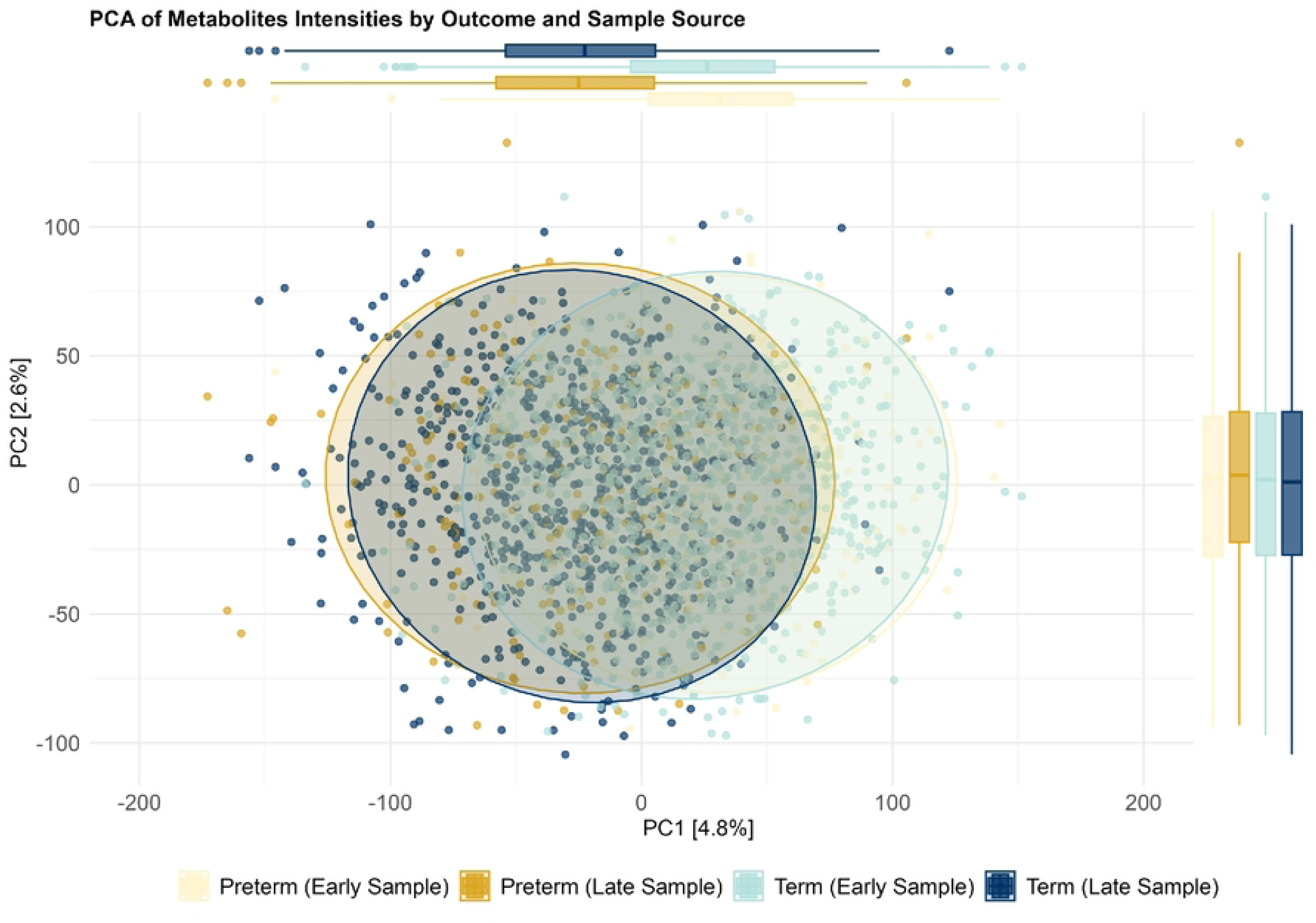
Principal Component Analysis of Maternal Metabolomic Profiles by Gestational Stage and Birth Outcome. PCA of maternal blood metabolomic data shows distinct clustering between early and late pregnancy samples, indicating metabolic shifts across gestation. Within each time point, term and spontaneous preterm birth (sPTB) samples overlap substantially. Permutational Multivariate Analysis of Variance (PERMANOVA) results (p = 0.455, R² = 0.0002) suggest that pregnancy outcome accounts for minimal variance in overall metabolomic profiles.

### Metabolite-Wise Association Analyses

To identify metabolomic associations with spontaneous preterm birth (sPTB), logistic regression analyses were performed on early and late pregnancy samples, as well as on the rate of metabolite change across gestation. The results are visualized in Fig 3.

**Fig 3:**
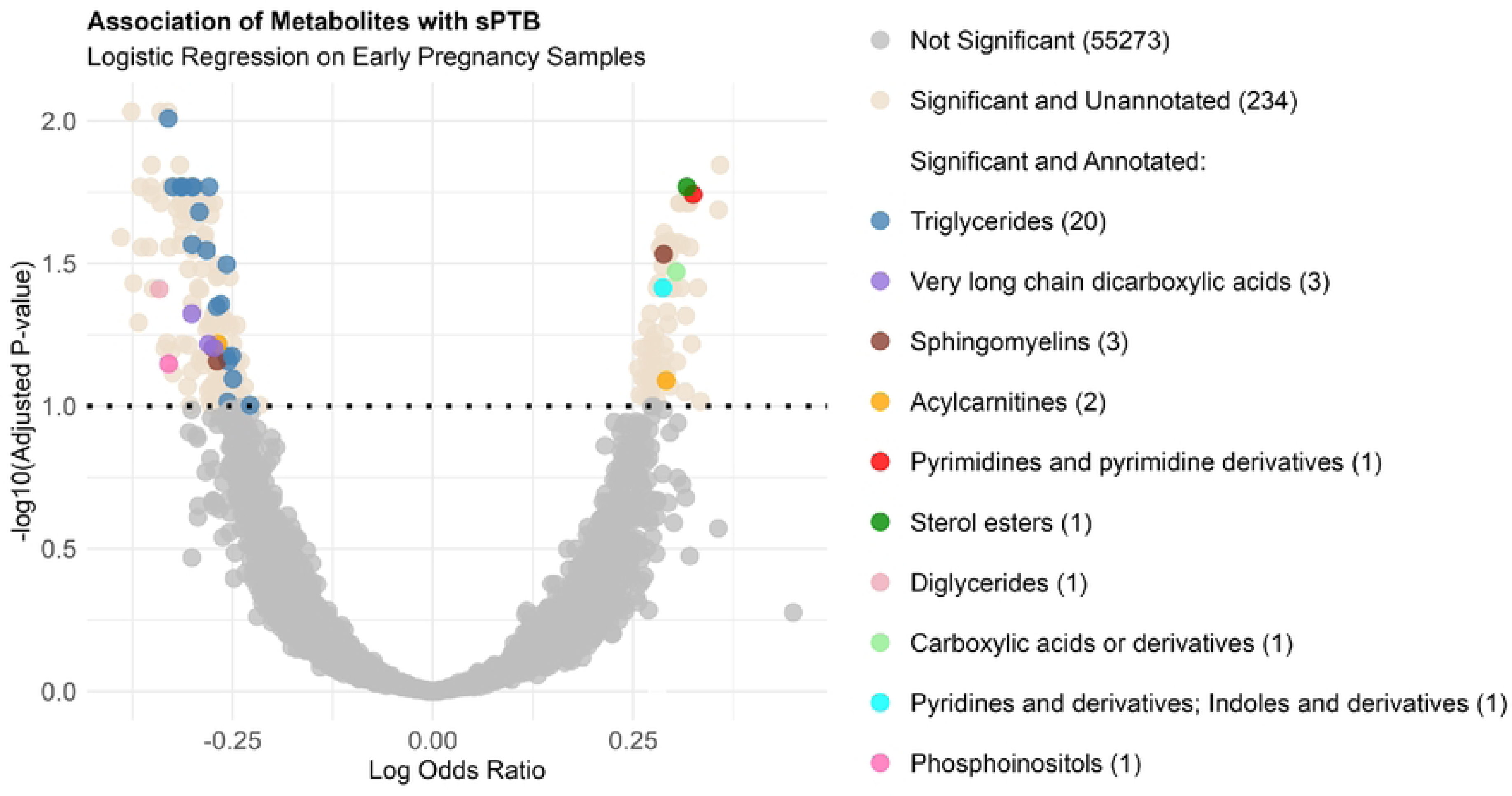
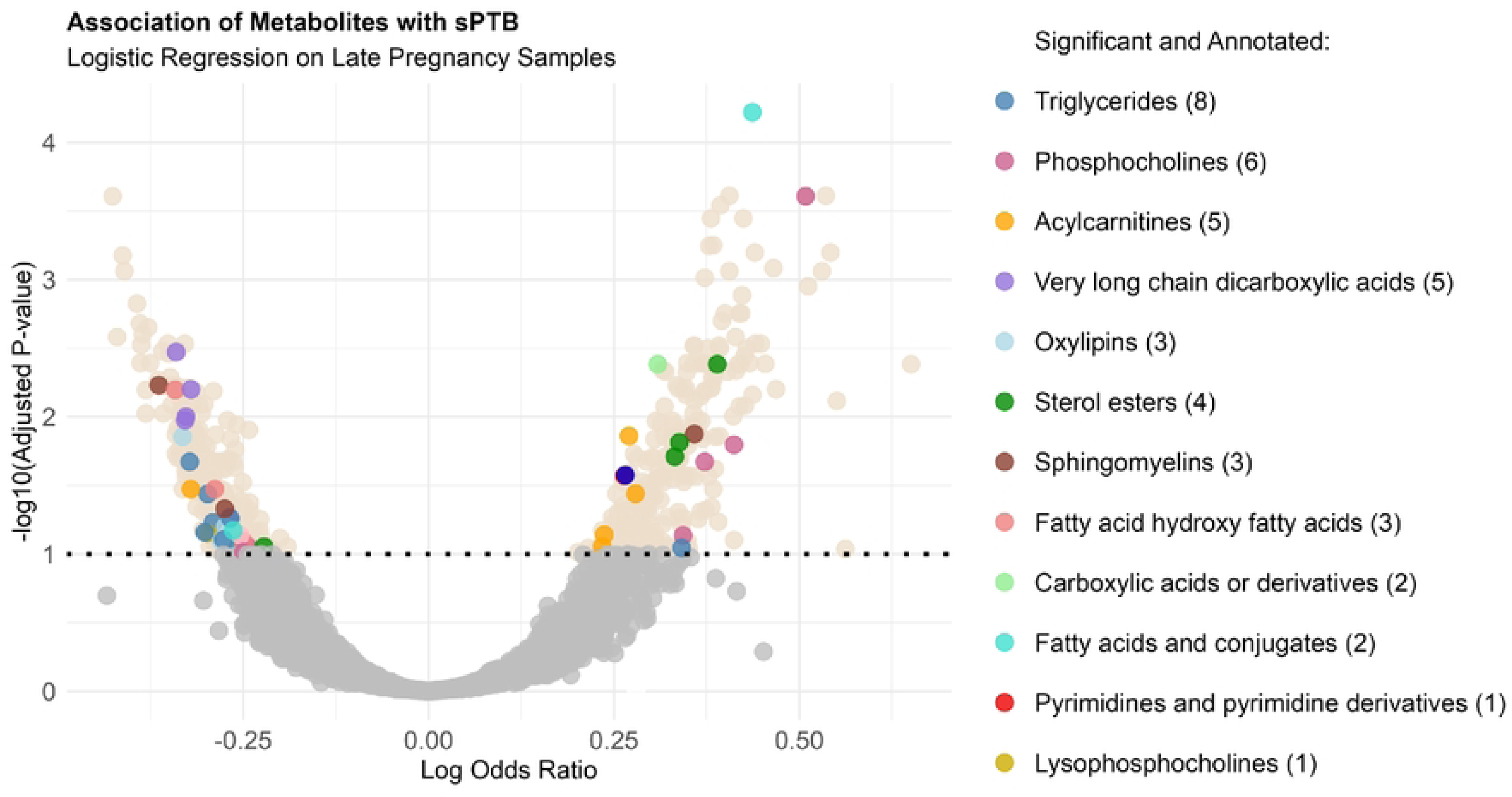
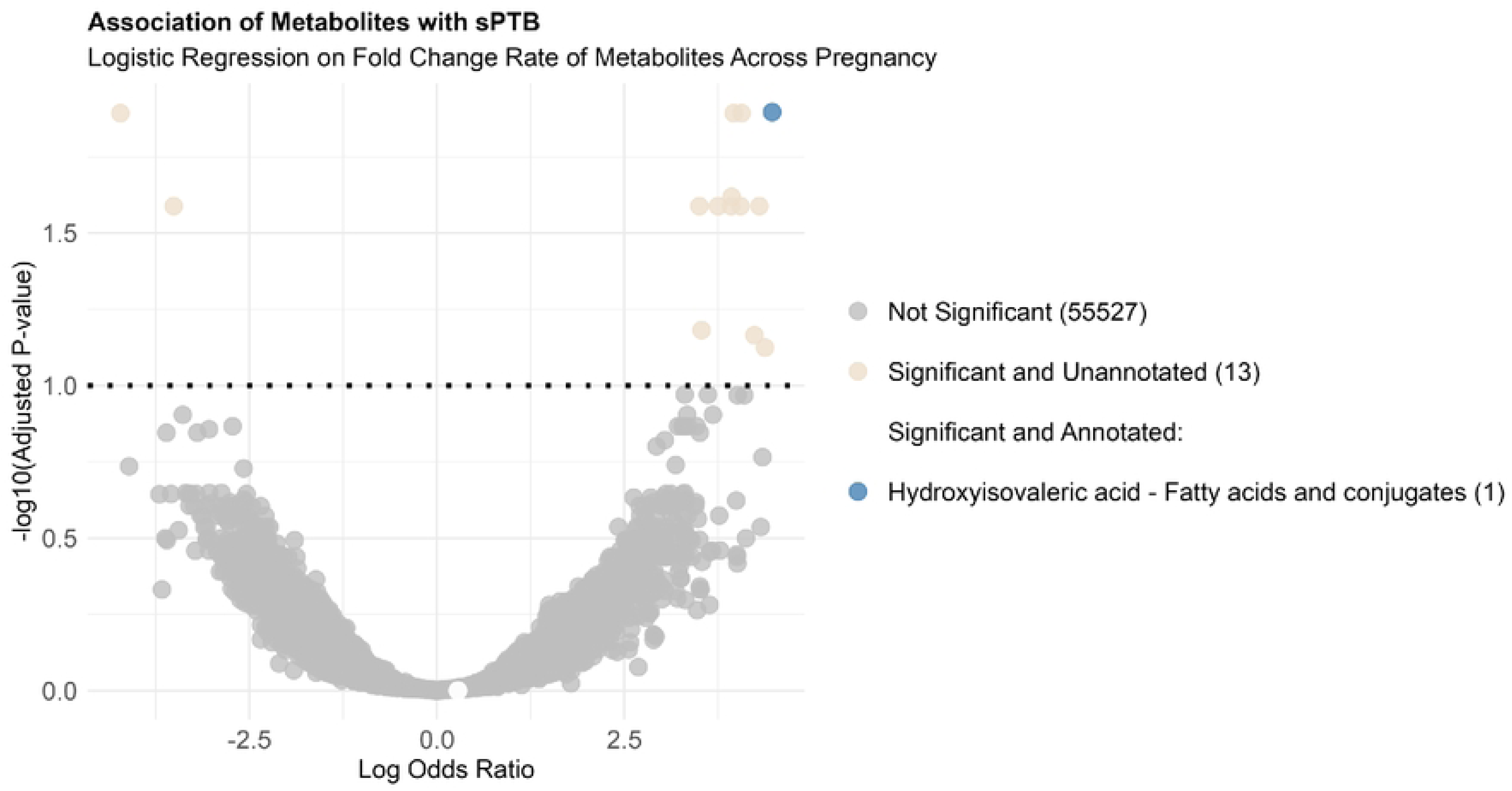
Volcano Plots of Metabolites Associated with Spontaneous Preterm Birth (sPTB) Volcano plots illustrating differentially expressed metabolites between sPTB cases and controls in (A) early pregnancy (< 20 weeks GA), (B) late pregnancy (≥ 20 weeks GA), and (C) metabolite fold change rates across pregnancy using two time-point paired samples. Each plot displays the log odds ratios from logistic regression models against the –log₁₀ of Benjamini-Hochberg adjusted p-values. Non-significant metabolites (FDR ≥ 0.1) are shown in grey; significant but non-annotated metabolites in beige; and significant annotated metabolites are color-coded by compound class.

Logistic regression analysis identified 268 metabolites in early pregnancy significantly associated with sPTB (FDR < 0.1), with 76 elevated and 192 decreased in cases; 34 of these were successfully annotated (Fig 3A). In late pregnancy, 617 metabolites were significantly associated with sPTB (FDR < 0.1), including 287 elevated and 330 decreased metabolites; 45 were annotated (Fig 3B). Longitudinal analysis revealed 14 metabolites whose rate of change during pregnancy was significantly associated with sPTB after adjusting for maternal age and early pregnancy expression levels (FDR < 0.1). Of these, 12 showed increasing trends and 2 decreasing trends in sPTB cases. Only one metabolite—Hydroxyisovaleric acid—was successfully annotated (Fig 3C).

Forest plots in Fig 4 display the direction and magnitude of association for each annotated metabolite with corresponding confidence intervals, aiding interpretation of key biomarker patterns across gestation. In early (< 20 weeks GA) pregnancy (Fig 4A), Dihydrothymine and 20:1 Cholesterol Ester were among the metabolites most associated with increased odds of sPTB, while TG(54:5) and TG(57:7) exhibited the greatest reductions in sPTB cases. In late (≥ 20 weeks GA) pregnancy (Fig 4B), PC(40:7:02) and Hydroxyisovaleric acid among the metabolites most positively associated with sPTB, whereas TG(57:5) and VLCDCA_C32H5204 demonstrated strong negative associations.

**Fig 4:**
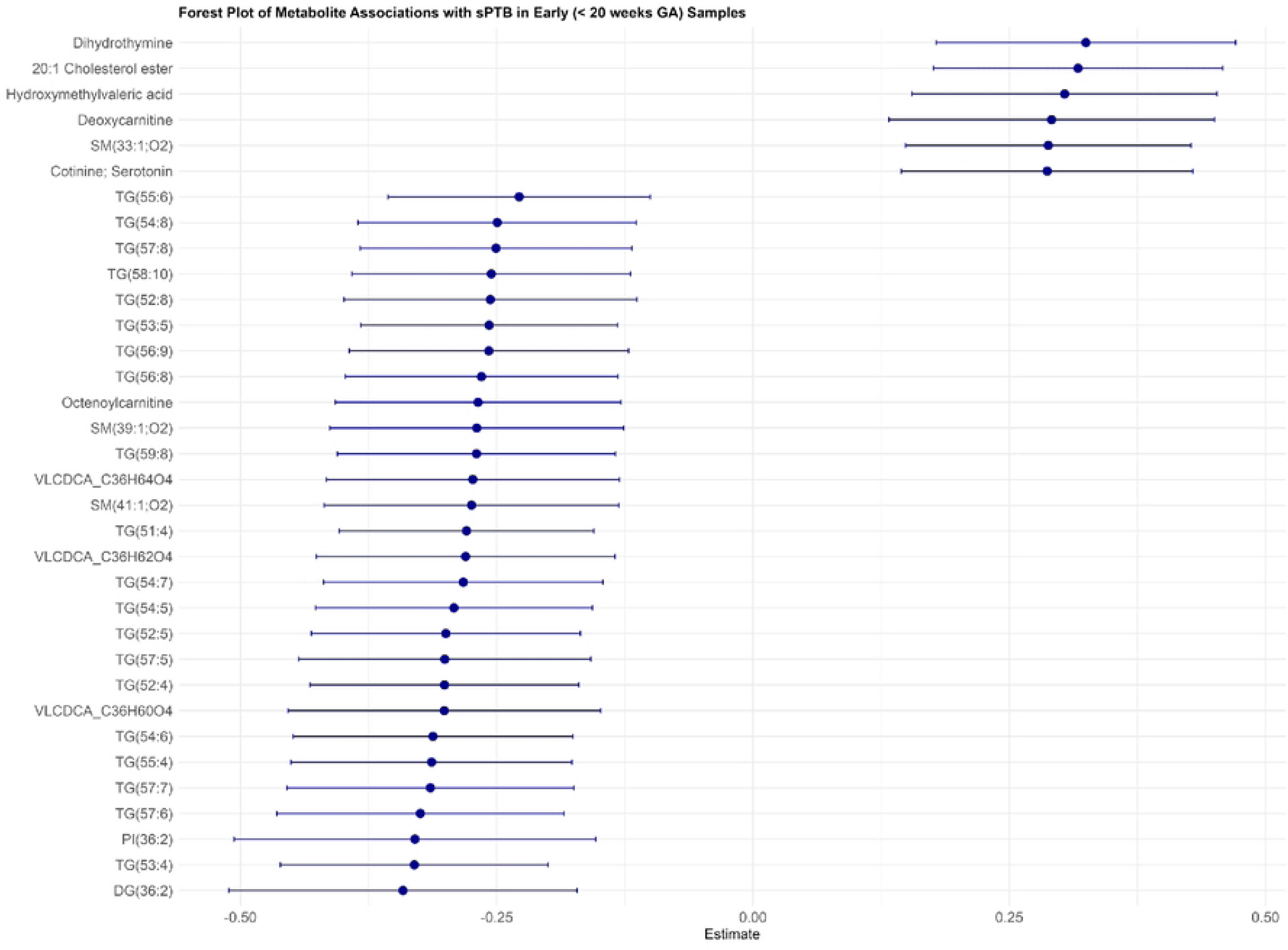
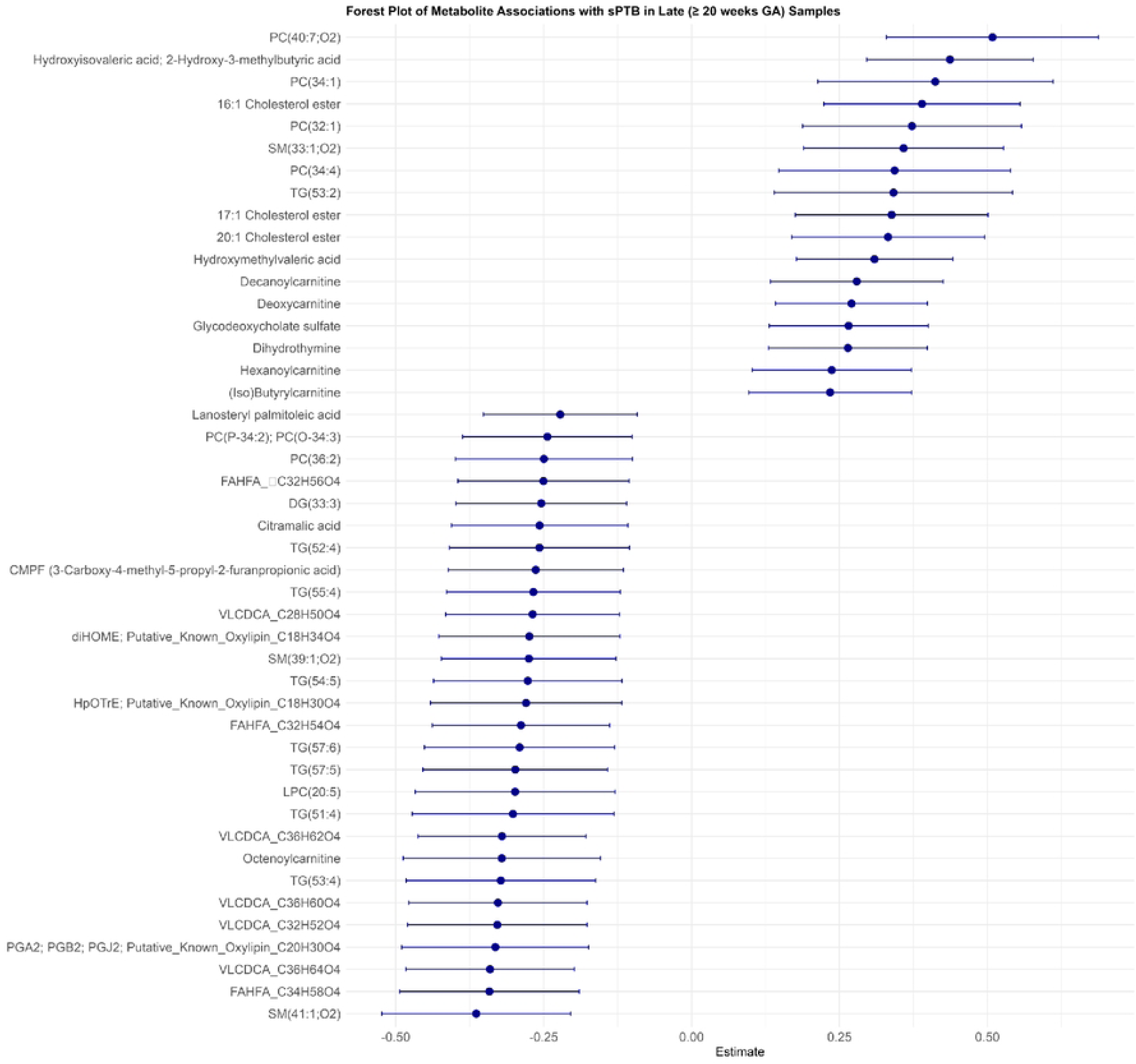
Forest Plots of Annotated Metabolites Associated with sPTB in Early and Late Pregnancy. Forest plots showing adjusted log odds ratios and 95% confidence intervals for annotated metabolites significantly associated with sPTB in (A) early (< 20 weeks GA) and (B) late (≥ 20 weeks GA) pregnancy samples. Results are based on logistic regression adjusted for maternal age.

The results from the logistic regression models for the metabolites that were not annotated with any form of identification are listed in S1 Fig, S2 Fig, and S3 Fig, along with their rLC-MS mass-to-charge ratios and retention times.

### Pathway Enrichment Analysis

Fig 5 presents the normalized enrichment scores (NES) for the top ten enriched compound classes using FGSEA for early and late pregnancy samples. For samples from early pregnancy, metabolites class triglycerides were significantly downregulated (FDR < 0.1) in sPTB group. On the other hand, the compound classes lysophosphocholines, bile acids and conjugates were upregulated. In late pregnancy samples, the compound class very long-chain dicarboxylic acids was significantly downregulated in sPTB (FDR < 0.1). Additionally, acylcarnitines and fatty acids and conjugates exhibited upregulation in sPTB, although these changes were not statistically significant. The observed differences between early and late samples reflect dynamic metabolic changes across gestation, emphasizing distinct compound class trends in sPTB.

**Fig 5:**
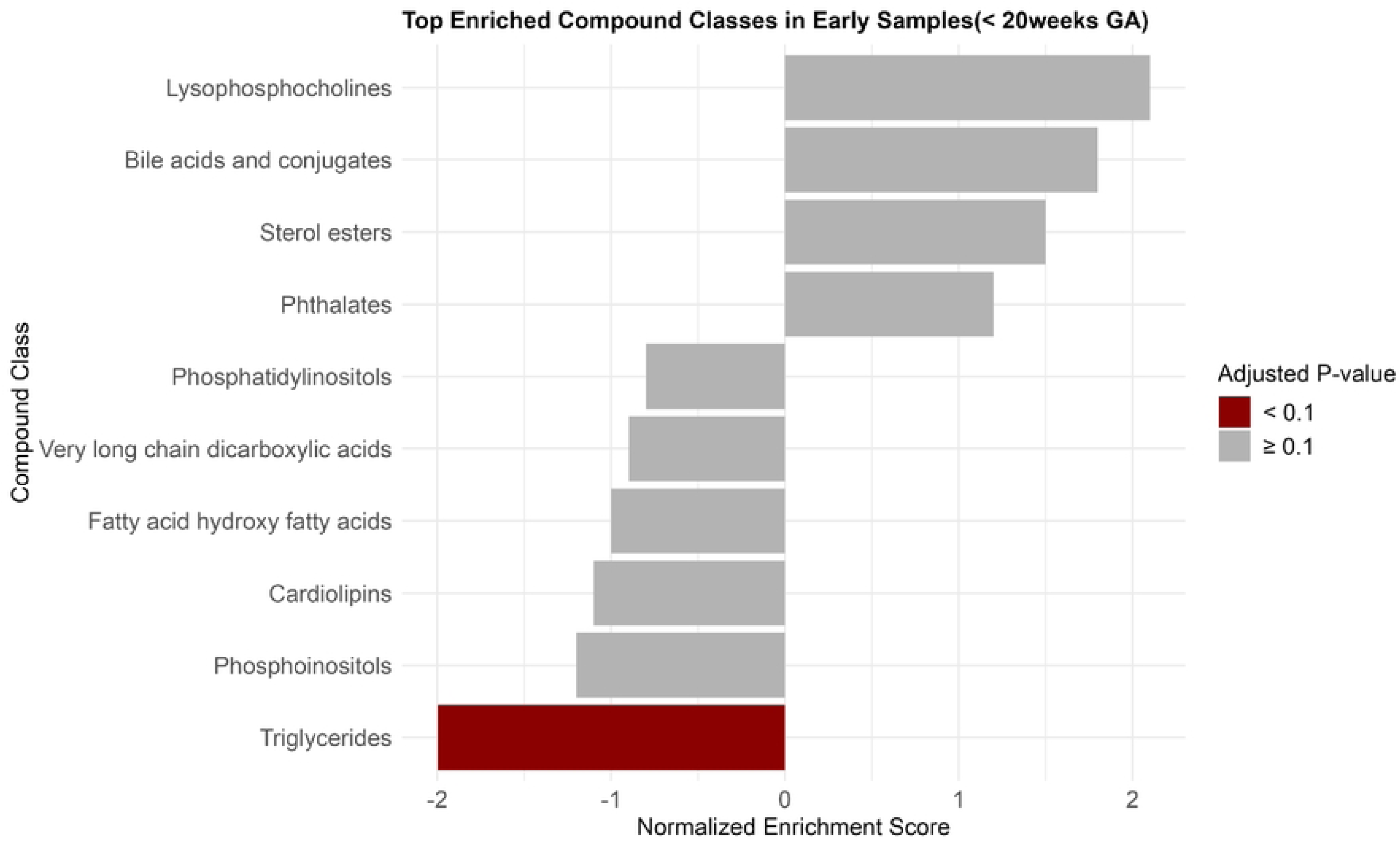
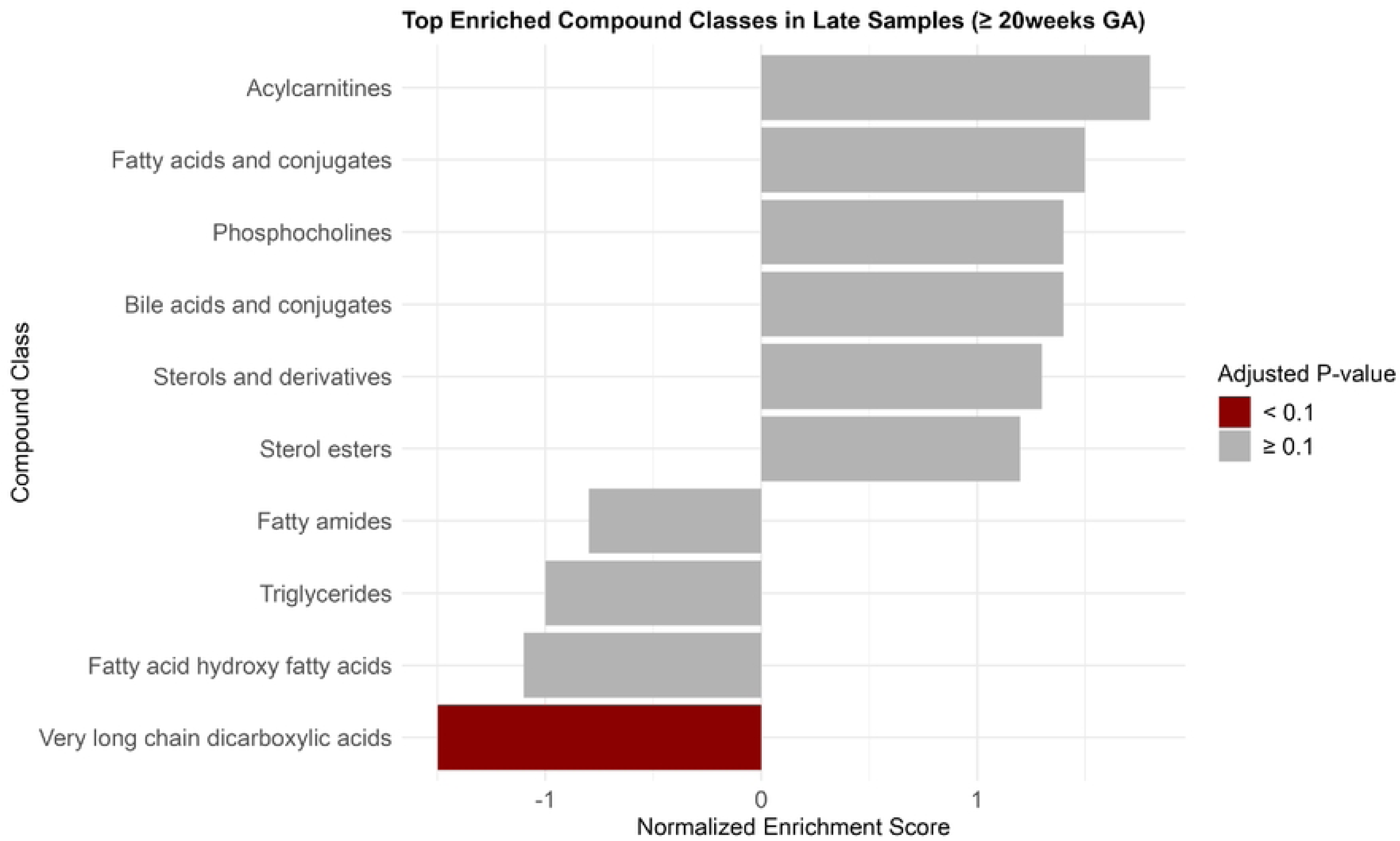
Enriched Compound Classes Associated with sPTB in Early and Late Pregnancy Blood Samples. Bar plots display the top enriched compound classes in (A) early and (B) late pregnancy blood samples associated with sPTB. Bars represent normalized enrichment scores; color indicates statistical significance based on adjusted p-values (FDR < 0.1 in red and ≥ 0.1 in gray)

## Discussion

This is a nested case-control analysis examining metabolomic alternations associated with sPTB within a large population-based cohort of pregnant women in Bangladesh. Blood plasma samples were collected at two distinct gestational time points—early and late pregnancy—to comprehensively characterize metabolic profiles. Using an untargeted metabolomics approach and a dual time-point strategy, we identified biomarkers specific to early and late stages of pregnancy, as well as metabolites whose rate of change across gestation—based on paired samples—was significantly associated with sPTB, highlighting both time-specific and dynamic metabolic indicators of risk, providing new insights into the biological mechanisms underlying sPTB. To our knowledge, this study represents one of the first focused on the application of untargeted metabolomics and its analysis in such a large cohort of pregnant women from a low- and middle-income country (LMIC) with a high burden of sPTB, like Bangladesh.

The most striking findings were the associations between lipid metabolites and sPTB. Twenty TGs were significantly downregulated in early pregnancy and seven in late pregnancy, whereas only TG (53:2) was upregulated in association with sPTB. Diglyceride (DG (36:2)) was also downregulated in early pregnancy. Additionally, sphingomyelins (SM), a class of membrane lipids, were significantly altered, with SM (41:1; O2) and SM (39:1; O2) downregulated and SM (33:1; O2) upregulated in sPTB. Other notable lipid alterations included disruptions in very long-chain carboxylic acids (VLCDA), phosphocholines (PC), lysophosphocholines (LPC), sterol esters, acylcarnitines, and olxylipins, all of which may contribute to impaired lipid metabolism, inflammation, and oxidative stress pathways in sPTB. These findings suggest that maternal metabolic disruptions, potentially influenced by factors such as insufficient nutrition status, low BMI and socioeconomic challenges, may contribute to sPTB risk.

Several studies have previously linked lipid levels and lipid metabolites to preterm birth. A U.S.-based study found that women delivering before 34 weeks had slower increases in TG levels by 21 weeks compared to those with term deliveries, aligning with our findings (27). Conversely, a meta-analysis of nine cohorts and five case-control studies (1,466 cases, 11,296 controls) suggested that elevated TG levels might increase sPTB risk (28). However, most studies in this review were conducted in high-income countries where pregnant women have different nutritional status and exposures compared to those in Bangladesh. Similarly, previous research has identified altered sphingomyelin metabolism in sPTB, with some studies reporting elevated SM levels in preterm cases while others found specific SM species to be downregulated (29–31). Beyond TGs and SMs, disruptions in phosphocholines (PCs), sterol esters, LPCs, acylcarnitines, and hydroxyisovaleric acid have also been linked to sPTB, with our study revealing differential expression patterns across gestation, highlighting the dynamic nature of lipid metabolism during pregnancy (32,33). By focusing on a Bangladesh cohort with distinct socioeconomic and environmental exposures, our study provides a complementary perspective to existing research.

Another important class of membrane lipids, phosphocholines (PC), were significantly associated with sPTB in our late pregnancy samples. Of the six significant, two were downregulated while four were upregulated in sPTB cases. Both increased and decreased levels of phosphocholines were found to be associated with sPTB in different studies (34,35). A phosphocholine-containing compound, heptadecanoyl-2-hydroxy-sn-glycero-3-phosphocholine has been established as a complementary predictor of preeclampsia, and so sPTB (33,36). Phosphocholine disruption may compromise cell membrane integrity, triggering pro-inflammatory cytokine release, which promotes cervical ripening, uterine contractions, and spontaneous preterm labor through inflammatory and oxidative stress pathways (37–39). We also found lysophosphocholine (LPC (20:5)) to be significantly lower in sPTB cases. Consistent with our findings, prior research has demonstrated a positive association between lysophosphatidylcholine (LPC) levels and birth weight. Furthermore, intrauterine growth restriction—a major contributor to low birth weight—has been strongly linked to spontaneous preterm birth (sPTB), thereby reinforcing the relevance of LPC in the pathophysiology of sPTB. (40–42). Our study also found decreased phosphoinositol levels in early sPTB samples, consistent with prior research linking low inositol in amniotic fluid to preterm birth. Inositol, essential for cell survival, signaling, and pulmonary surfactant production, may contribute to sPTB risk through disrupted cellular functions and impaired fetal lung development (43–45).

Acylcarnitines, esters of fatty acids and L-carnitine, are essential for β-oxidation and energy metabolism, often serving as markers for metabolic disorders (46). A published study reported differential acylcarnitine expression in preterm birth cases (47). Hexanoylcarnitine and decanoylcarnitine, elevated in our sPTB cases, align with findings linking these markers to preeclampsia, a known cause of sPTB (33,48). However, our findings of elevated deoxycarnitine and isobutyrylcarnitine and decreased octanoylcarnitine differ from previous studies, possibly due to variations in maternal acylcarnitines across gestation (49). Increased deoxycarnitine suggests bacterial vaginosis, and elevated isobutyrylcarnitine is linked to gestational diabetes, both associated with sPTB (33,50,51). Our novel discovery of decreased octanoylcarnitine in both gestational periods may indicate impaired β-oxidation of medium-chain fatty acids, potentially affecting fetal growth (52).

Another fatty acid-based class of compounds, oxylipins which are derived from polyunsaturated fatty acids visa oxidation, also showed three species being significantly downregulated in late samples of sPTB. Oxylipins are linked to gestational age at birth and play crucial roles in pregnancy by influencing immune activation, endothelial cell function, inflammation resolution, and coagulation (53,54). Specifically, decreased levels of diHOME and prostaglandin PGJ2 determined in our study aligns with previous literature as they show to have a protective effect on spontaneous preterm birth (54). A fatty acid conjugate, 3-carboxy-4-methyl-5-propyl-2-furanpropanoic acid (CMPF) is known to be elevated in the plasma of humans with gestational diabetes mellitus, a cause of sPTB, and in our study, late samples showed significant elevation of CMPF in sPTB (55,56). Our findings highlight the critical role of lipid and fatty acid metabolism in pregnancy. We also determined hydroxy fatty acids to be significantly negatively associated with sPTB in late samples which could be established as novel markers as well.

Beyond lipid-related metabolites, we identified significant associations between other metabolites and sPTB, including dihydrothymine, bile acids, and their conjugates. Dihydrothymine, a pyrimidine degradation product, was significantly elevated in both early and late pregnancy sPTB cases, suggesting potential disruptions in nucleotide metabolism and oxidative stress pathways (57,58). To our knowledge, this is the first study to report an association between dihydrothymine and sPTB. Altered bile acid metabolism may indicate underlying hepatic dysfunction, which has been implicated in adverse pregnancy outcomes, including preterm birth (59–61).

We also found that cotinine and serotonin were significantly upregulated in early pregnancy sPTB cases. Cotinine, a nicotine metabolite and biomarker of tobacco exposure, was elevated in sPTB cases, suggesting possible maternal nicotine exposure, likely from consumption of betel nut/leaf with tobacco, which is common among rural Bangladeshi women, and passive smoking exposure (62). Nicotine exposure can impair placental function, fetal development, and induce inflammation, increasing sPTB risk (63–66). Serotonin, a biogenic amine and neurotransmitter, plays a key role in vascular regulation, placental function, and inflammatory pathways. Altered serotonin levels have been associated with maternal stress and antidepressant use, which may influence sPTB risk through effects on uterine contractility (67,68). The role of betel quid exposure and selective serotonin reuptake inhibitors (SSRIs) in sPTB warrants further investigation in our cohort.

Beyond the unique study population, our study has two other major strengths. First, the use of untargeted metabolomics provided a comprehensive, unbiased approach for detecting a wide range of metabolites across diverse metabolic classes. Among the 55,000 metabolites detected, 650 were identified as significantly associated with sPTB (to be confirmed). Although many of these metabolites lack matched HMDB IDs, limiting immediate pathway analysis, future annotation efforts will enable deeper mechanistic insights. Second, our dual time-point sampling strategy—analyzing both early and late gestation samples—allowed us to capture the temporal dynamics of metabolic changes. Notably, we identified 14 metabolites whose rate of change during pregnancy was associated with sPTB, emphasizing the importance of longitudinal metabolomic profiling in understanding preterm birth risk.

Our study has some limitations and major differences as other studies in the literature. First, we adjusted only for maternal age and did not account for other covariates, which may contribute to differences between our findings and those in the literature. Given that metabolites may lie on the causal pathway between socioeconomic, physiological, or environmental factors and sPTB, adjusting for these variables could lead to overadjustment and underestimation of the effect of the metabolites under investigation. Second, while our dual time-point sampling provides insight into metabolic changes during pregnancy, the absence of denser longitudinal follow-up, limits our ability to assess metabolic trajectories at a more granular level. Additionally, potential selection bias due to the rural cohort and relatively small sample size is another limitation. Future studies incorporating broader epidemiological, environmental, and socioeconomic factors, along with mediation analyses and extended longitudinal sampling, will be necessary to further validate these findings and improve our understanding of metabolomic changes in sPTB.

## Conclusion

This study provides novel insights into the metabolomic alterations associated with sPTB in a rural Bangladeshi population. Using an untargeted metabolomics approach, we identified significant disruptions in lipid metabolism, particularly in triglycerides, sphingomyelins, and phosphocholines, suggesting potential mechanistic links to poor nutritional status and metabolic dysregulation. Additionally, associations with bile acids, dihydrothymine, cotinine, and serotonin highlight the influence of both metabolic and environmental factors on sPTB risk. Given the high burden of sPTB in LMICs, future research should integrate longitudinal metabolomic profiling, socioeconomic determinants, and maternal exposures to identify predictive biomarkers and inform targeted maternal health interventions aimed at reducing preterm birth.

## Acknowledgments

We gratefully acknowledge the study participants in Bangladesh—including the women, children, and their families—for their invaluable time, biological samples, and data. Appreciation is also due to the AMANHI study staff at host institutions in Bangladesh and partner organizations abroad, as well as to the Government of Bangladesh, including the Ministry of Health and relevant district and local authorities, for their ethical oversight, logistical support, and commitment to facilitating the smooth execution of the study.

## Data Availability

The dataset used and analyzed for this manuscript will be available from the corresponding author on request.

## Supplementary Information

**S1 Table: Logistic Regression Results for Unannotated Metabolites Associated with sPTB in Early (< 20 weeks GA) Pregnancy Samples.** This table presents the results of logistic regression analyses assessing the association between metabolite intensities in early pregnancy samples and spontaneous preterm birth (sPTB). Only unannotated metabolites are included here, with their corresponding mass-to-charge (m/z) ratios and retention times (RT) provided to facilitate future identification or validation efforts.

**S2 Table: Logistic Regression Results for Unannotated Metabolites Associated with sPTB in Late (≥ 20 weeks GA) Pregnancy Samples.** This table presents the results of logistic regression analyses assessing the association between metabolite intensities in late pregnancy samples and spontaneous preterm birth (sPTB). Only unannotated metabolites are included here, with their corresponding mass-to-charge (m/z) ratios and retention times (RT) provided to facilitate future identification or validation efforts.

**S3 Table: Logistic Regression Results for Unannotated Metabolites Associated with sPTB in Fold Change Rate Model Involving Paired Samples from Two Time-points.** This table presents the results of logistic regression analyses assessing the association between metabolite fold change rates across pregnancy and spontaneous preterm birth (sPTB). Only unannotated metabolites are included here, with their corresponding mass-to-charge (m/z) ratios and retention times (RT) provided to facilitate future identification or validation efforts.

## Notes

### Competing Interest Statement

The authors have declared no competing interest.

### Funding Statement

This work was supported by the Bill & Melinda Gates Foundation (Grant No. INV-005276). In accordance with the Gates Foundation’s agreement with the Public Library of Science (PLOS), we understand that PLOS journals will publish manuscripts funded by the Gates Foundation without an article processing charge (APC). We respectfully request that this manuscript be considered under that arrangement.

